# Effects of central venous pressure trajectories on the risk of acute kidney injury in patients with severe sepsis

**DOI:** 10.1101/2025.06.01.25328758

**Authors:** Jia Chen, Xiao-Fan Mao, Han-Bing Wang, Xiao-Hong Lai

## Abstract

**Background:** Recent years, the trajectory models enable clinicians to explore longitudinal data. Previous studies had found that high central venous pressure (CVP) exposure significantly increase acute kidney injury (AKI). However, no literature had revealed the association between CVP trajectories and AKI occurrence in septic patients. This study aims to investigate the dynamic evolution of CVP and its association with AKI development.

**Methods:** This was a retrospective observational cohort study using data collected from the Medical Information Mart for Intensive Care IV (MIMIC-IV) , an open-source clinical database. Severe septic adult patients with CVP measurements and subsequently diagnosed with AKI were included. Group based trajectory model (GBTM) was used to derive heterogeneous CVP trajectories and analyze the AKI rate in each trajectory. Univariable and multivariate logistic regression was used to detect the association between variables and outcome.

**Results:** A total of 1372 patients were enrolled in our study. Three trajectories were identified in the results: the consistently moderate and stable CVP class (Class 1), the class with an elevation trend and then mild decline but stay persistently high level (Class 2), and the transient mild decline precedes sustained increase class (Class 3). These classes showed differences in AKI rate. Class 1 had the lowest rate (63.15%), Class 2 the highest (92.10%), and Class 3 was intermediate (86.05%) (p<0.001). After model adjustment, Class 2 continued to possess the highest risk (p<0.001). The baseline characteristics of age (OR=1.02, 95%CI=1.01–1.02) and BMI (OR=1.06, 95%CI=1.04–1.07) are risk factors of AKI.

**Conclusions:** Patients with persistently high CVP and those with transient mild decline precedes sustained increase of CVP had a greater risk of developing AKI than patients with consistently moderate and stable level. The different trajectories of CVP could provide risk markers for adverse events of AKI.

## Background

Severe sepsis is a lethal condition where infection triggers pathological immune signaling cascades and may culminate in multi-system organ failure[1]. Globally, there are more than 30 million sepsis patients each year[2]. In spite of the advances in rapid recognition and resuscitation, sepsis remains to be a major cause of morbidity and mortality worldwide[3]. The 2017 Global Burden of Disease study estimated 48.9 million global sepsis cases with 22.5% mortality, contributing to 20% of all deaths worldwide[4–6]. The impact of sepsis is not only on in-hospital survival but extends into post-discharge quality of life[7]. The systemic inflammatory response in severe sepsis often results in multi-organ dysfunction, with kidney impairment representing the most frequently and earliest[8–11]. The medical academic community has made sustained efforts toward early identification and management of sepsis induced renal damage.

Central venous pressure (CVP) is an important indicator in critically ill patients in addition to other basic vital signs[12]. Previous studies have identified CVP as the exclusive hemodynamic parameter linked to acute kidney injury (AKI) onset[13]. High CVP exposure may significantly increase AKI, which indirectly increase mortality[14, 15]. These findings highlight the need for further investigation into the relationship between CVP and AKI. Recently, there were several researches investigated the relationship of serum creatinine or urine-output trajectories with AKI development [16, 17]. However, no literature had revealed the association between CVP trajectories and AKI in septic patients. The variation tendency of CVP could be crucial. Herein, we sought to characterize the association of CVP trajectories and AKI development in severe septic patients.

## Methods

### Data sources

The Medical Information Mart for Intensive Care IV (MIMIC IV) is an anonymous, and publicly available database which contains data from over 50,000 admissions from 2008 to 2019 at the Beth Israel Deaconess Medical Center. The MIMIC-IV database was used to gather data for the study (https://mimic-iv.mit.edu/) on April 10,2025. One of our members had obtained a CITI certificate and did the data extraction. Patient authorization and ethical approval were waived. The authors had no access to information that could identify individual participants during or after data collection.

### Study population

Our cohort composed by patients with severe sepsis (ICD-code R652, R6520, R6521) who developed AKI after 24h of intensive care unit (ICU) admission and had CVP measurements of the first day in ICU. If the patient had two times of ICU admission, only the first one was taken. The CVP value should be measured at every 6h after admission and presented as average in each period. The exclusion criteria were: age <18, AKI was noted before ICU admission, patient with CVP counts less than 3 times of the 4 periods and outlier value more than 2, variable values missing more than 20%, patients who received kidney replacement, patients died in 72h after ICU admission. If missing values or outliers were present only once for a patient, the average of the other three times was used for imputation. Kdigo-stages≥1 was defined as AKI development based on the KDIGO guidelines [18]. The other covariates of demographic features, including age, sex, Body Mass Index (BMI) and ethnicity along with the comorbidities, including hypertension, diabetic and coronary heart disease (CHD) of the patients were also extracted.

### Statistical analysis

Categorical variables and presented as proportion (%). Shapiro-Wilk and Levene tests were performed for continuous variables. Mean (standard deviation) or medians (interquartile range) were used for presenting continuous variables. The Mann– Whitney test was utilized for the comparison between the AKI group and the non-AKI group. Chi-square test or Fisher’s exact test was used for the categorical variables’ comparison between classes with adjustment for Bonferroni method to mitigate Type I error inflation. Kruskal-Wallis was used for continuous variables. P < 0.05 was considered to be statistically significant. The missing data were filled by multiple imputation or average. Group based trajectory model (GBTM) was used to analyze the longitudinal data and explore the heterogeneity[19]. The optimal trajectory model was picked out according to the following criteria: (1) the lowest Bayesian information criterion (BIC) and Akaike information criterion (AIC); (2) the highest Log-Likelihood (Loglik); (3) higher average posterior probabilities ( Avepp ) for each trajectory group [20]. The association between CVP trajectory and AKI was explored by multivariate logistic regression to calculate odds ratios (OR) with 95% confidence intervals (CI). Univariate Logistic Regression was used to detect the association between each individual variable and AKI. The forest plot was used for this visualization. Statistical analyses were performed using R (version 4.4.2).

## Results

A total of 1372 adults including 776 men and 596 women aged 18 to 91 years (65.96 ±15.81years) were enrolled in this research. The detailed flow chart for enrollment and exclusion is provided in Fig 1. The average BMI was 29.62kg/m^2^ with a standard deviation of 7.79 kg/m^2^. There were 931 White patients (67.86%),101 Black (7.36%),44 Asian (3.21%) and 296 other races (21.57%) in this cohort. The total ICU-acquired-AKI rate of patients with severe sepsis was 65.74%. Except for gender, the other characteristics were statistic different between non-AKI group and AKI group (p<0.05). The characteristics of eligible patients between the two groups are shown in Table 1.

**Fig 1.**
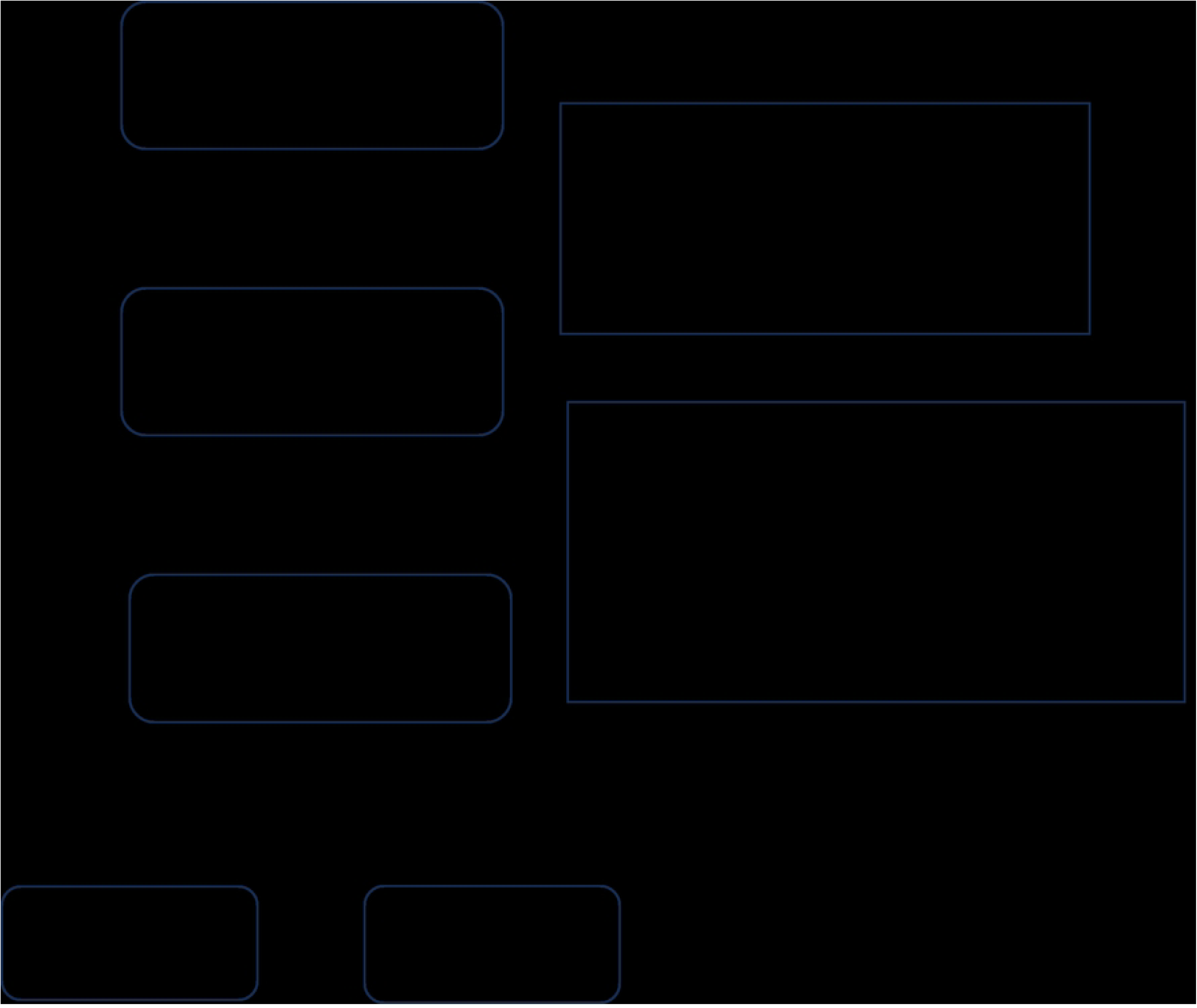
**The detailed flow chart for enrollment and exclusion**. ICU: intensive care unit; AKI: acute kidney injury; CVP: central venous pressure; GBTM: Group based trajectory model.

**Table 1.**
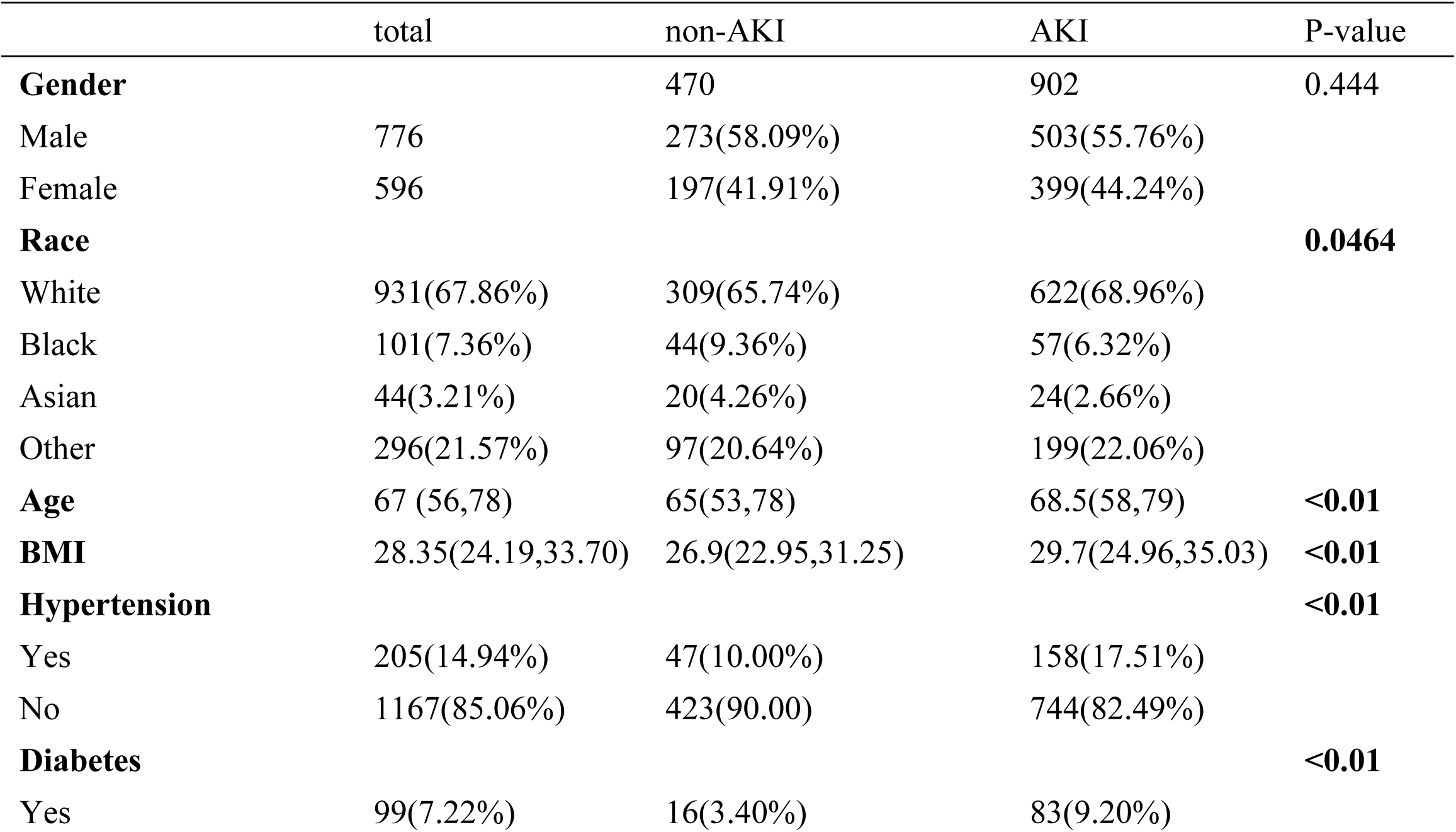
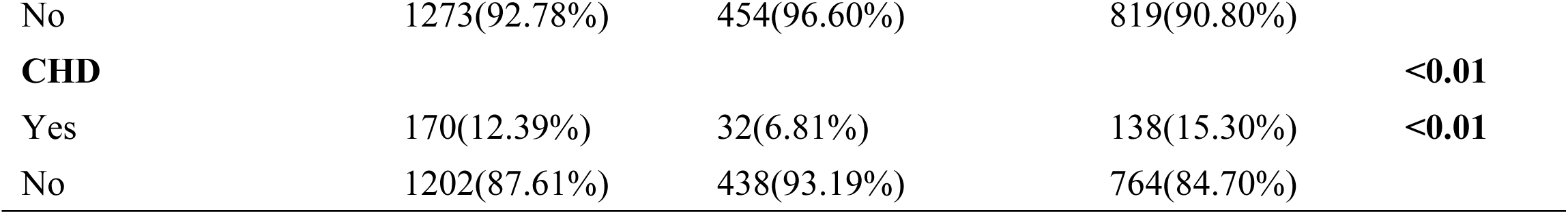
Baseline characteristics between the non-AKI group and AKI group. AKI: acute kidney injury; CHD: Coronary heart disease; Data are presented as counts (percentage) or median (25%,75% percentile) due to the different types of variables.

|  | total | non-AKI | AKI | P-value |
| --- | --- | --- | --- | --- |
| **Gender** |  | 470 | 902 | 0.444 |
| Male | 776 | 273(58.09%) | 503(55.76%) |  |
| Female | 596 | 197(41.91%) | 399(44.24%) |  |
| **Race** |  |  |  | **0.0464** |
| White | 931(67.86%) | 309(65.74%) | 622(68.96%) |  |
| Black | 101(7.36%) | 44(9.36%) | 57(6.32%) |  |
| Asian | 44(3.21%) | 20(4.26%) | 24(2.66%) |  |
| Other | 296(21.57%) | 97(20.64%) | 199(22.06%) |  |
| **Age** | 67 (56,78) | 65(53,78) | 68.5(58,79) | **<0.01** |
| **BMI** | 28.35(24.19,33.70) | 26.9(22.95,31.25) | 29.7(24.96,35.03) | **<0.01** |
| **Hypertension** |  |  |  | **<0.01** |
| Yes | 205(14.94%) | 47(10.00%) | 158(17.51%) |  |
| No | 1167(85.06%) | 423(90.00) | 744(82.49%) |  |
| **Diabetes** |  |  |  | **<0.01** |
| Yes | 99(7.22%) | 16(3.40%) | 83(9.20%) |  |
| No | 1273(92.78%) | 454(96.60%) | 819(90.80%) |  |
| **CHD** |  |  |  | **<0.01** |
| Yes | 170(12.39%) | 32(6.81%) | 138(15.30%) | **<0.01** |
| No | 1202(87.61%) | 438(93.19%) | 764(84.70%) |  |

The Group based trajectory model (GBTM) was used to calculate the loglikelihood, AIC, BIC and Avepp of different sectionalization. We systematically constructed models beginning with a 1 class and progressively extended to 4 classes. The results are presented in Table 2. There was little difference in BIC between 3 classes and 4 classes classification, but BIC tended to be selected conservatively to avoid overfitting. The Loglik/AIC improvement was not significant from 3 classes to 4 classes. Besides, the 4 classes group had 2 classes’ proportion lower than 5%. Therefore the 3 classes group model was picked out. Among these 3 classes, the AKI rate ranged from 63.15% to 92.10%. These trajectories showed differences in CVP baseline and variation tendency (Fig 2).

**Fig 2.**
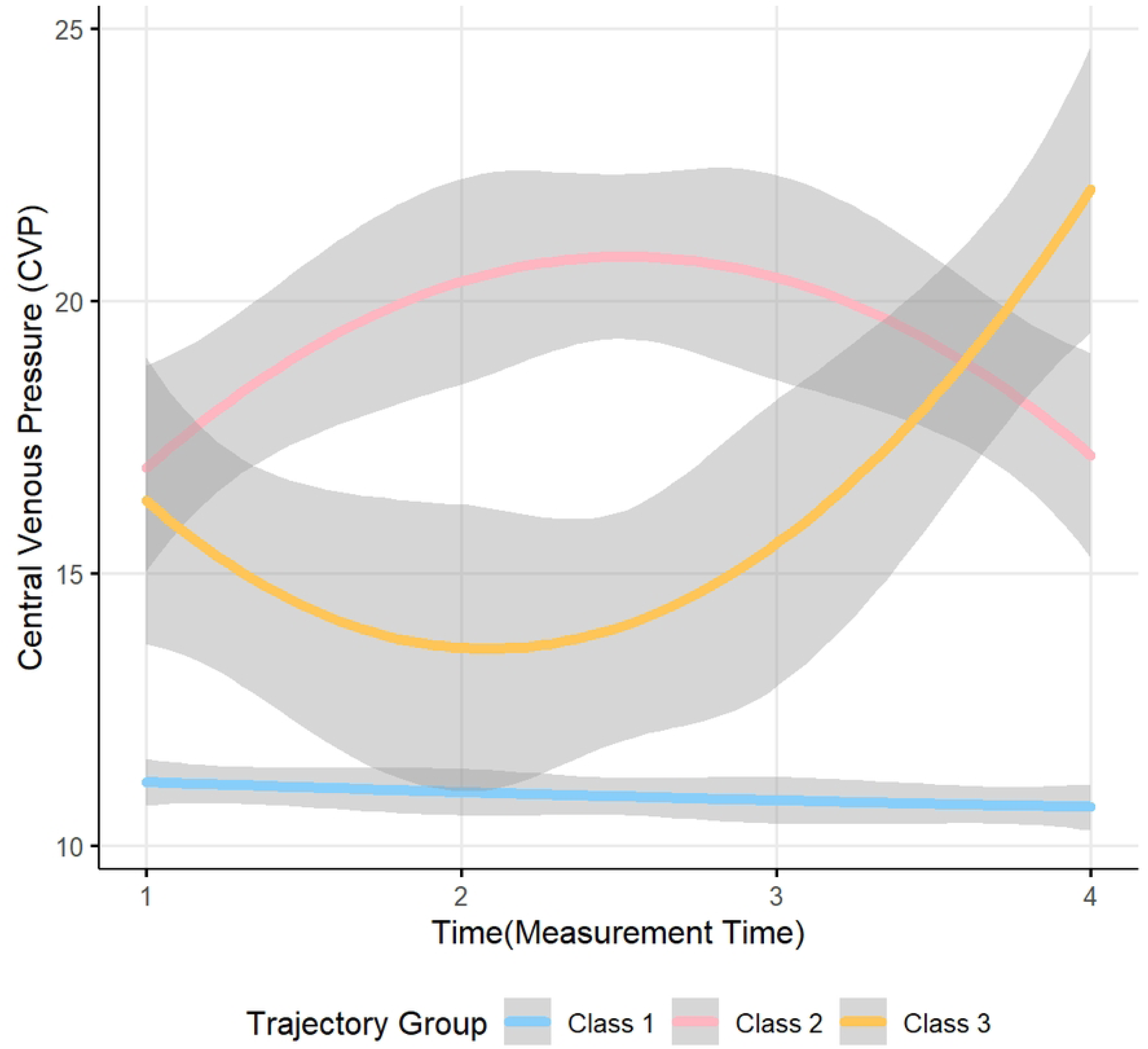
**The CVP trajectory of different classes**. Confidence intervals are shown in the shaded areas. The measurement time was every 6h on the first day in ICU.

**Table 2.** Trajectory model parameters. AIC: Akaike information criteria; BIC: Bayesian information criteria; Aevpp: Average posterior probability

| Class |  |  |  | 1 | 2 | 3 | 4 | 1 | 2 | 3 | 4 |
| --- | --- | --- | --- | --- | --- | --- | --- | --- | --- | --- | --- |
|  | Loglik | AIC | BIC | Aevpp(%) |  |  |  | proportion(%) |  |  |  |
| 1 | -16614.62 | 33235.24 | 33250.92 | 100 |  |  |  | 100 |  |  |  |
| 2 | -14976.85 | 29975.7 | 30033.18 | 96.27 | 80.90 |  |  | 92.29 | 7.71 |  |  |
| 3 | -14909.83 | 29849.65 | 29928.03 | 95.82 | 78.90 | 79.53 |  | 89.47 | 6.64 | 3.89 |  |
| 4 | -14859.27 | 29756.54 | 29855.82 | 79.00 | 95.20 | 82.15 | 85.63 | 7.28 | 88.50 | 2.70 | 1.52 |

*Class 1. CVP trajectory in this class showed a stable low level (<12cmH2O) from baseline after ICU admission and followed by a mild decline*.

*Class 2. CVP trajectory in this class started with a much higher baseline (>15cmH2O) and soon reached its peak, then a slight drop back showed up but still higher than 15 cmH2O*.

*Class 3. CVP trajectory in this class also started with a high baseline but a minor decrease emerged quickly. After that, sustained increase occurred and CVP value reached a surprisingly high level (>20cmH2O) at the end of the day*.

The baseline characteristics of age and BMI differed among these three classes (p<0.05). In addition, the AKI rate also showed significant differences among the 3 classes (p<0.001). Further tests showed this ratio between Class 2 and Class 1 was significant (p<0.001), as well as in Class 3 and Class 1 (p=0.011). But there was no significant difference between Class 2 and Class 3 (p=0.98). The comparison of the baseline characteristics and AKI rate by the different trajectory classes are showed in Table 3.

**Table 3.** Baseline characteristics and AKI rate by the different trajectory classes. AKI: acute kidney injury; CHD: Coronary heart disease; Data are presented as counts (percentage) or median (25%,75% percentile) due to the different types of variables.

|  | Class1 | Class2 | Class3 | P-value |
| --- | --- | --- | --- | --- |
| **Gender** |  |  |  | 0.3352 |
| Male | 696(56.13%) | 51(57.30%) | 29(67.4%) |  |
| Female | 544(43.87%) | 38(42.70%) | 14(32.6%) |  |
| **Race** |  |  |  | 0.121 |
| White | 847(68.31%) | 63(70.79%) | 21(43.84%) |  |
| Black | 94(7.58%) | 3(3.37%) | 4(9.3%) |  |
| Asian | 39(3.15%) | 3(3.37%) | 2(4.65%) |  |
| Other | 260(20.97%) | 20(22.47%) | 16(37.21%) |  |
| **Age** | 68(37,88) | 65(35.4,86.6) | 62(29,84.8) | **0.0417** |
| **BMI** | 28.2(19.3,43.9) | 30.7(21.6,48.6) | 30.7(22.7,42.4) | **<0.001** |
| **Hypertension** |  |  |  | 0.6293 |
| Yes | 189(15.24%) | 11(12.36%) | 5(11.63%) |  |
| No | 1051(84.76%) | 78(87.64%) | 38(88.37%) |  |
| **Diabetes** |  |  |  | 0.7865 |
| Yes | 91(7.34%) | 6(6.74%) | 2(4.65%) |  |
| No | 1149(92.66%) | 83(93.26%) | 41(95.35%) |  |
| **CHD** |  |  |  | 0.2834 |
| Yes | 156(12.58%) | 10(11.24%) | 4(9.3%) |  |
| No | 1084(87.42%) | 79(88.76%) | 39(90.7%) |  |
| **AKI** |  |  |  | **<0.001** |
| Yes | 783(63.15%) | 82(92.1%) | 37(86.05%) |  |
| No | 457(36.85%) | 7(7.9%) | 6(13.95%) |  |

Univariable logistic regression was performed to evaluate the association of individual variables with the primary outcome, and generate a forest plot to visualize the odds ratio (OR) with 95% confidence interval (95%CI). The correlation between age and AKI was significant, along with BMI. Each 1-year increase in age (OR=1.02, 95%CI=1.01–1.02, p<0.001) and each 1-unit increase in BMI (OR=1.06, 95%CI=1.04– 1.07, p<0.001) were independent risk factors for AKI. However none of the other baseline characteristics showed significant association with AKI development (p>0.05). The results are presented in Fig 3.

**Fig 3.**
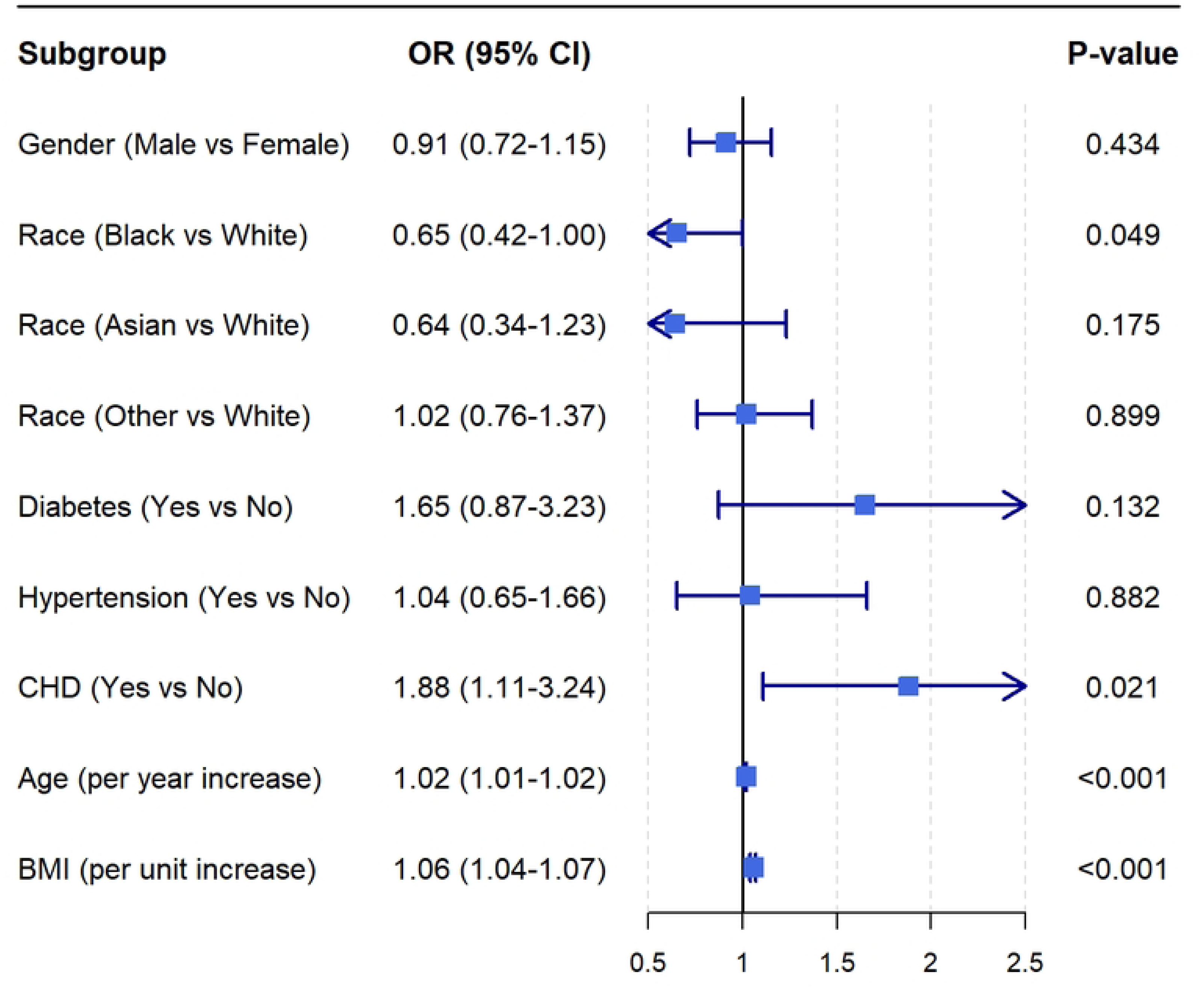
**Subgroup analysis of the association with different variable and the AKI rate**. Subsequently, we performed a multivariate logistic regression to further compare the risks of different classes relative to Class 1. After adjusting for some demographic factors (age, gender, race, BMI), the odds ratio (OR) was 4.35 of Class 2 (95%CI=3.10-5.33, p<0.001) and 3.77 of Class 3 (95%CI=1.66-10.15, p=0.004). Both indicated significantly higher risk compared to the reference group (Class 1) as in model 1. Then after adjusting for all the factors including the comorbidities, the risk was still significant as in Model 2 (Class 2: OR=4.54, 95%CI=3.18–5.82, p<0.001; Class 3: OR=3.99, 95%CI=1.76–10.74, p=0.004). The results of multivariate logistic regression analysis were presented in Table 4.

**Table 4.** Comparison of different classes with Class 1 after adjusting for multiple factors. Model 1: adjust for age, gender, race and BMI; Model 2: adjust for age, gender, race, BMI, diabetes, hypertension and coronary heart disease. Ref is short for reference.

| Variable | Model 1 |  |  |  | Model 2 |  |  |  |
| --- | --- | --- | --- | --- | --- | --- | --- | --- |
|  | OR | CI low | CI high | P | OR | CI low | CI high | P |
| Class1 | Ref | Ref | Ref | Ref | Ref | Ref | Ref | Ref |
| Class2 | 4.35 | 3.10 | 5.33 | **<0.001** | 4.54 | 3.18 | 5.82 | **<0.001** |
| Class3 | 3.77 | 1.66 | 10.15 | **0.004** | 3.99 | 1.76 | 10.74 | **0.002** |

## Discussion

AKI is one of the most frequent organ dysfunctions associated with severe sepsis and may lead to increased morbidity and mortality[21]. Previous studies have suggested the use of interleukins (ILs) or other molecular biomarkers for early prediction of AKI[22–24]. However, these indicators are not always cost-efficient and easily accessible in clinical practice. In this study, we primarily investigated the association between early CVP trajectories and AKI occurrence during the first 24 hours in ICU among patients with severe sepsis. We derived three heterogenous trajectory patterns and found that the persistently high CVP trajectory (even though followed by a slight downtrend) was associated with the highest incidence of AKI. These findings are in accordance with Runlu Sun’s viewpoint who found that a 1 mmHg increase in CVP increases the odds of AKI in critically adult patients[15]. Our research also found that for patients with relative elevated CVP initially and further rises to more than 20mmHg within a short period, the incidence of AKI rates was also higher. CVP measurement alone has little effect on the outcome of septic AKI[25]. Our study gave a potential link between dynamic changes in CVP and the development of AKI.

Maintain an appropriate CVP level can help to keep patients hemodynamically stable, ensure renal perfusion, and prevent further ischemic injury due to ongoing renal dysfunction[26]. Our results revealed that the CVP trajectory may serve as an easily accessible early warning marker for AKI occurrence. A longer duration of high CVP was related to the poor prognosis[27]. As elevated CVP particularly impairs renal hemodynamics and contributes to AKI by increasing renal ‘afterload’ and reducing renal perfusion pressure[28]. From our perspective, closer monitoring of CVP and assessment of its fluctuation could be essential in severe septic patients. It is necessary for the clinicians to pay attention to those septic patients whose CVP stay at a continuous high level resemble Class 2 or increase rapidly resemble Class 3 at the early phase. Prompt intervention needs to be implemented for those critically unstable patients.

Our research also revealed that the ICU-acquired-AKI rate of patients with severe sepsis in MIMIC IV database was up to 65.74%. Meanwhile each 1-year increase in age and each 1-unit increase in BMI were independently associated with elevated AKI risk. Simultaneously patients in the AKI group had a higher proportion of comorbidities as well. These findings are consistent with previous studies[29–31]. For those patients are more vulnerable to the onslaught of inflammatory response. Previous studies have identified racial disparities in AKI incidence[32], which were not consistent with our findings. The divergence could potentially stem from heterogeneity in the different studied populations.

We recognize there were several limitations in our investigation. First of all, since this study was conducted retrospectively at a single institution, future prospective trials across multiple centers are necessary to confirm our findings. Secondly the cohort was lack of laboratory parameters before ICU admission which may provide more subtle information. Thirdly CVP is the result of a combination of multiple factors, trajectory model can’t elucidate the underlying mechanisms. Future multicenter studies with larger population are needed to substantiate these observations and enhance their clinical applicability.

## Conclusions

CVP trajectories in patients with severe sepsis within the first 24h in ICU were heterogeneous. These trajectories identified patients with disparate clinical characteristics and risk for development of AKI. Patients with persistently high CVP and those with relatively high level followed by a rapid ascending trend had a greater risk of developing AKI than patients with consistently moderate and stable CVP. The different trajectories of a CVP may serve as risk markers for adverse events of AKI.

## Data Availability

Data can be shared publicly after acceptance

## Acknowledgements

We would like to express our gratitude to Chao-Jin Chen, Ji-Rong Yang and Yu-Heng Luo from the Third Affiliated Hospital of Sun Yat-sen University for their valuable assistance in this study.

